# Chronic Adaptive Deep Brain Stimulation in Parkinson’s Disease: ADAPT-START Findings and Programming Principles

**DOI:** 10.1101/2025.09.25.25336275

**Authors:** Simona Cascino, Fabrizio Luiso, Elena Contaldi, Laura Caffi, Chiara Palmisano, Gianni Pezzoli, Ioannis Ugo Isaias, Salvatore Bonvegna

## Abstract

Deep brain stimulation (DBS) is an established treatment for advanced Parkinson’s disease (PD), yet conventional DBS (cDBS) may provide suboptimal symptom control and can induce adverse effects, particularly on gait. Adaptive DBS (aDBS), which dynamically adjusts stimulation amplitude in response to subthalamic beta oscillatory activity, offers the potential for superior outcomes; however, its clinical benefits and programming strategies remain incompletely defined. Between January and April 2025, we consecutively offered the opportunity to test aDBS with the dual threshold algorithm to the first 20 PD patients with chronic cDBS and a Percept family neurostimulator who attended scheduled visits at our center. Nine were eligible and tested the aDBS mode. The primary reasons for exclusion or delayed programming were the presence of signal artifacts, absence of a distinct alpha-beta peak, and clinically optimized stimulation settings that were incompatible with aDBS. Of nine eligible patients, by July 2025, five entered chronic aDBS, one reverted to cDBS by preference, and three remained in the optimization phase. In the five patients on chronic aDBS, unblinded MDS-UPDRS III assessment showed an average 35% greater motor improvement compared with cDBS. Gait outcomes improved most consistently, with an average 40% reduction in Freezing of Gait Questionnaire scores. These preliminary findings suggest that the dual threshold aDBS mode with the Percept system may provide clinical advantages over cDBS and is preferred by most eligible patients, although technical challenges and programming demands currently limit broader clinical implementation.

## INTRODUCTION

Deep brain stimulation (DBS) of the subthalamic nucleus (STN) is a safe and highly effective neurosurgical treatment, and a well-established, evidence-based therapy for patients with Parkinson’s disease (PD)^1^. The treatment consists of brain-implanted electrodes connected to a pulse generator (IPG), usually placed in the subclavicular region. Conventional DBS (cDBS) delivers continuous stimulation with fixed parameters, which may be suboptimal for managing axial symptoms (e.g., gait)^2^, as well as motor and non-motor fluctuations. These symptom fluctuations are invariably present in advanced PD and are associated with disease progression and residual dopaminergic medication^3^. They are further modulated by everyday activities; for example, cognitive tasks may exacerbate tremor or trigger freezing of gait^4^.

In recent years, technological advances have enabled the use of chronically implanted electrodes to record real-time local field potentials (LFP), which are extracellular electrical potentials reflecting the dynamics and synchronized activity of nearby neuronal and glial populations^5^. In PD, elevated beta-band (13–35 Hz) oscillations recorded from the STN have been associated with the severity of akinetic-rigid symptoms^6^ and with fluctuations related to dopaminergic medication^7^; they are generally suppressed by STN-DBS^8,9^.

In this context, subthalamic LFP recordings may optimize cDBS programming by mapping the location of beta power to guide the selection of optimal stimulation contacts^10,11^ Such recordings have also been explored as input signals to enable timely adjustment of the stimulation delivered (adaptive DBS, aDBS). In principle, aDBS could improve motor fluctuations, allowing for better integration of DBS with dopaminergic medication^12^. Additionally, aDBS paradigms might enable automatic adjustments of brain stimulation based on motor and non-motor activities in daily life, which are also encoded in subthalamic oscillatory activity^13–16^.

Two commercially available devices currently offering aDBS are the AlphaDBS (Newronika SpA), which employs a “linear proportion” algorithm, and the Percept family neurostimulators (Medtronic Inc.), which operates in “single threshold” and “dual threshold” modes^17^. Feasibility studies have been successfully conducted, with clinical outcomes from both short-^4,18^ and long-term^12,19^ follow-ups forthcoming. However, real-world evidence regarding the chronic use of aDBS remains largely anecdotal, with very little information available on programming strategies and troubleshooting^20,21,22^.

In this study, we present our initial experience and workflow for programming patients with the Percept device using aDBS and the dual threshold algorithm. We highlight key programming strategies as well as potential pitfalls associated with this novel stimulation mode.

## METHODS

Between January 1 and April 30, 2025, we consecutively enrolled patients with idiopathic PD from the Parkinson Institute of Milan – ASST Gaetano Pini-CTO, who had been chronically treated with STN-DBS using a Percept device, to test aDBS with dual threshold mode (Figure 1), a feature now implemented in all Percept family devices and available to patients based on clinical indications. Aiming to offer aDBS to all patients as a new, potentially superior treatment, we applied the single inclusion criteria of having of stable and optimized chronic stimulation parameters with cDBS for the preceding four months. Optimal stimulation contacts had been clinically determined in the past for cDBS through a monopolar review^23^, aided by *BrainSense Surveys* and image-guided programming, and remained unchanged throughout the study period. To ensure a consistent comparison between cDBS and aDBS, oral medication regimens were maintained without modification.

**Figure 1.**
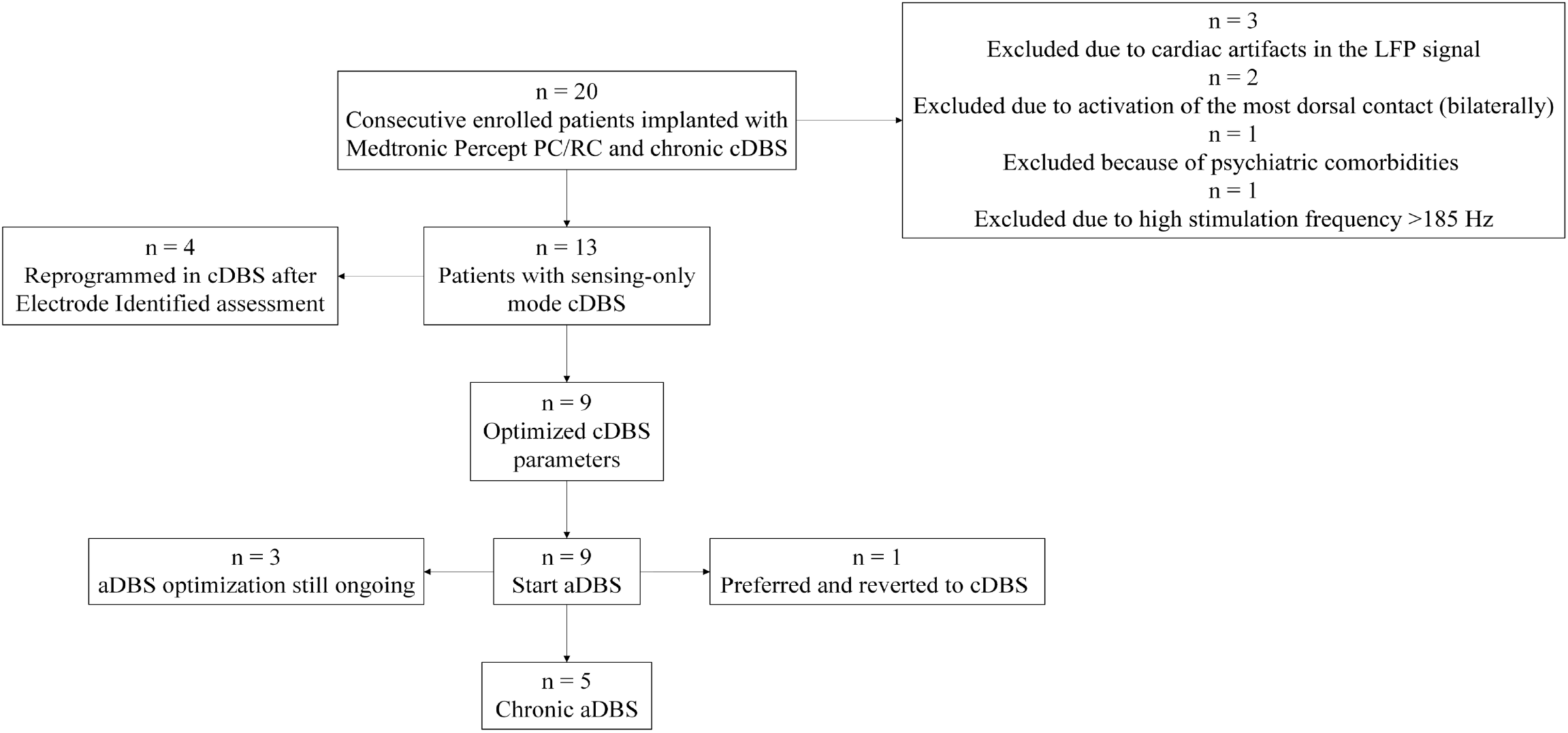
Patient timeline.

In this study, we initially aimed to follow the programming workflow proposed in the ADAPT-PD trial^24^, but some modifications were required during implementation. The ADAPT-PD trial is a prospective, single-blind, randomized crossover clinical study aimed at demonstrating the safety and the effectiveness of chronic single and dual threshold aDBS modes with the Percept family neurostimulators. We applied only the dual threshold mode for all patients. This algorithm gradually modulates the stimulation amplitude between two limits identified by the clinician, increasing it over a 2.5-minute period when beta power exceeds an upper LFP threshold and decreasing it over a 5-minute period when beta power remains below a lower LFP threshold. Changes in current amplitude occur in steps of 0.1 mA for the Percept PC (Primary Cell) and 0.01 mA for the RC (Rechargeable Cell) if the beta power stays outside the threshold range for more than 1.2 seconds. When beta power returns within the two thresholds, the stimulation amplitude is held constant at the last reached value. The monitored beta power is calculated within a patient-specific frequency range of 5 Hz (fixed frequency range), centered around the most prominent alpha-beta peak.

In this initial phase of aDBS use, our programming approach followed two guiding principles: (i) minimizing patient exposure to adverse effects (such as troublesome dyskinesias or OFF states); and (ii) limiting the number of additional programming visits for aDBS setup to three, with one extra visit permitted. If aDBS could not be optimised within these visits, aDBS programming was deferred to routine follow-up.

No formal statistical analysis of the results was conducted because of the limited number of patients who reached the 1-month follow-up. Instead, we assessed the Minimal Clinically Important Difference (MCID^25^), as clinical relevance was considered particularly informative at this early stage of aDBS.

For the first **SET-UP visit** (Table 1, Box 1), patients arrived at the clinic with chronic cDBS and overnight withdrawal of antiparkinsonian medication (cDBS-ON/Meds-OFF). We performed the following:

**Table 1:** Detailed description of each programming visit.

| **Box 1 – SET-UP VISIT**<br>**(cDBS-ON/Meds-OFF or wearing OFF)** |  |
| --- | --- |
| **Clinical assessment** | • MDS-Unified Parkinson's Disease Rating Scale (MDS-UPDRS) parts I–IV<br>• Freezing of Gait Questionnaire (FOG-Q)<br>• Parkinson's Disease Questionnaire (PDQ-39) |
| **Device and stimulation check** | • Battery life check<br>• Impedance check<br>• Retrieve chronic cDBS stimulation settings. *In the case of a new activation, perform a clinical monopolar review (aided by BrainSense Surveys)* |
| **BrainSense Surveys**<br>**(Electrode Identifier and Electrode Survey)** | • Define and review alpha-beta peaks |
| **BrainSense Setup**<br>**Signal Test and activation of**<br>**Sensing-Only mode** | • Review and compare alpha-beta peak automatically identified with results from previous BrainSense Surveys and clinical programming. If the alpha-beta peak is not detected, it must be identified manually<br>• Activate cDBS with Sensing Only mode for 3-5 days recordings |
| **BrainSense Streaming** | • Assess modulation of the alpha-beta power by increasing stepwise the stimulation from 0 to 3 mA (steps of 0.5 or 1 mA for 30 sec each). If no modulation is detected, change sensing contact pair |
| **Events** | • Enable home-based events (e.g., medication intake, bedtime and wake-up time, and, when applicable, motor OFF periods and dyskinesias) |
| **Box 2 – ACTIVATION VISIT**<br>**(cDBS-ON/Meds-OFF or wearing OFF)** |  |
| **Timeline** | • Check LFP power fluctuation for both hemispheres It shows 24h interval of LFP activity (10 min averaged bins). Particularly check also for awake/asleep changes |
| **Events LFP Snapshots**<br>**Timeline** | • Verify that the alpha-beta peak from Snapshots and spectral analysis (from Timeline) matches the chronic sensing alpha-beta peak. Check also for LFP magnitude changes across different type of events<br>– **If no matching:** Use the chronic alpha-beta peak, continue Sensing Only mode for another 3-5 days at home<br>– **If matching:** Proceed with LFP streaming (below) |
| **BrainSense Streaming** | • Identify lower and upper stimulation limit as follows: turn stimulation OFF bilaterally; clinically evaluate the patient and start BrainSense Streaming; record LFP for one minute in resting state; increase stimulation in 0.5 mA steps for one hemisphere at the time and monitor clinical response and LFP modulation (10 sec).<br>– **Lower stimulation limit:** minimum amplitude giving >40-50% clinical improvement over Stim-OFF;<br>– **Upper stimulation limit:** maximum effective amplitude before clinical side effects. This value needs to be confirmed in Meds-ON<br>Lower and upper stimulation limits should be tested separately for each hemisphere, followed by bilateral Stim-ON. These limits represent the minimum and maximum current that will be delivered by the IPG<br>• Set **Pause amplitude**, typically between the lower and upper stimulation limits, which is delivered when stimulation is paused (often matching conventional stimulation amplitude) |
| **Timeline** | • Based on the Timeline over multiple days, identify LFP thresholds:<br>– lower LFP threshold: 25% cut-off of beta power of wakeful chronic recordings<br>– upper LFP threshold: 75% cut-off of beta power of wakeful chronic recordings |
| **Clinical Response Testing** | • Monitor the patient, particularly during the first activation for 1-2 hours, requesting the patient to remain near the clinic |
| **Box 3 - FOLLOW-UP VISIT**<br>**(aDBS-ON/Meds-ON)** |  |
| **2-week follow-up** | • Interview and clinically evaluate the patient, review Timeline, LFP Chart and Events LFP Snapshots. If new symptoms arise (e.g., dyskinesias, OFF states) adjust alpha-beta power thresholds and secondarily amplitude limits (revisit the steps outlined in Box 2) |

i. Check battery level, impedance, and stimulation settings.
ii. Neurological evaluation using the Movement Disorder Society Unified Parkinson’s Disease Rating Scale (MDS-UPDRS) parts I–IV and the Montreal Cognitive Assessment (MoCA). Based on our previous experience showing gait improvement with aDBS^19^, we aimed to further investigate this aspect using the Freezing of Gait Questionnaire (FOG-Q)^26^. For a more comprehensive assessment of daily activities over time, patients also completed the Parkinson’s Disease Questionnaire (PDQ-39)^27^. (Table 2).
iii. Identification of optimal sensing contacts and alpha-beta peaks through the *BrainSense Surveys* and *BrainSense Setup*. The recordings were visually inspected, and contacts contaminated by cardiac or movement artifacts –particularly heel strikes observed during brief back-and-forth walking– were excluded.
iv. Live streaming (*BrainSense Streaming*) with incremental increase in stimulation amplitude (0.5 mA steps) to assess beta modulation and responsiveness to stimulation.

**Table 2:** Comparison of clinical scales and questionnaires in patients who successfully transitioned to aDBS mode after 1-month follow-up.

| **Clinical motor scales** | **cDBS-OFF /**<br>**Meds-OFF**<br>**median score**<br>**(range)** | **cDBS-ON /**<br>**Meds-OFF**<br>**median score**<br>**(range)** | **aDBS-ON /**<br>**Meds-OFF**<br>**median score**<br>**(range)** | **Changes in Median Score**<br>**aDBS-ON / Meds-OFF vs.**<br>**cDBS-ON / Meds-OFF** | **Changes in Median Score**<br>**cDBS-OFF / Meds-OFF vs.**<br>**cDBS-ON / Meds-OFF** | **Changes in Median Score**<br>**cDBS-OFF / Meds-OFF**<br>**vs. aDBS-ON / Meds-OFF** | **MCID**<br>**(n. of patients)**<br>**Imp. / Wors.** |
| --- | --- | --- | --- | --- | --- | --- | --- |
| MDS-UPDRS part III (max: 132) | 37 (33-58) | 23 (10-29) | 15 (6-18) | ↓ 8 (35%) | ↓ 14 (38%) | ↓ 22 (59%) | 4 / 0 |
| *Bradykinesia (items 3.4 - 3.8) (max: 40)* | 19 (10-21) | 10 (5-13) | 7 (1-10) | ↓ 3 (30%) | ↓ 9 (47%) | ↓ 12 (63%) | – |
| *Rigidity (item 3.3 (max: 20)* | 10 (2-12) | 4 (1-5) | 1 (0-4) | ↓ 3 (75%) | ↓ 6 (60%) | ↓ 9 (90%) | – |
| *Tremor (items 3.15-3.18) (max: 40)* | 4 (0-16) | 0 (0-4) | 0 (0-3) | – | ↓ 4 (100%) | ↓ 4 (100%) | – |
| *Axial symptoms & Gait (items 3.1, 3.2, 3.9 - 3.14) (max: 12)* | 10 (4-18) | 8 (2-11) | 4 (2-9) | ↓ 4 (50%) | ↓ 2 (20%) | ↓ 6 (60%) | – |
| **Clinical anamnestic scales and**<br>**questionnaires** | **cDBS-ON / Meds-ON**<br>**median score (range)** |  | **aDBS-ON / Meds-ON**<br>**median score (range)** |  | **Changes in Median Score**<br>**cDBS-ON / Meds-ON vs. aDBS-ON / Meds-ON** |  | **MCID**<br>**(n. of patients)**<br>**Imp. / Wors.** |
| MDS-UPDRS part I (max: 52) | 11 (7-13) |  | 9 (2-15) |  | ↓ 2 (18%) |  | 3 / 1 |
| MDS-UPDRS part II (max: 52) | 9 (4-11) |  | 8 (6-10) |  | ↓ 1 (11%) |  | 3* / 2 |
| MDS-UPDRS part IV (max: 24) | 2 (0-4) |  | 2 (0-7) |  | – |  | 2 / 2 |
| *Dyskinesia (items 4.1 - 4.2) (max: 4)* | 0 (0-2) |  | 0 (0-1) |  | – |  | – |
| *Motor OFF (items 4.3 - 4.6) (max: 16)* | 0 (0-3) |  | 1 (0-7) |  | ↑ 1 (100%) |  | – |
| FoG-Q (max: 24) | 5 (1-7) |  | 3 (2-8) |  | ↓ 2 (40%) |  | – |
| PDQ-39 (max: 100)** | 13 (7-29) |  | 18 (7-28) |  | ↑ 5 (28%) |  | – / 3 |
| *Mobility (max: 100)* | 20 (3-60) |  | 30 (3-40) |  | ↑ 10 (33%) |  | – / 2 |
| *Activity of Daily Living (max: 100)* | 13 (8-21) |  | 8 (4-38) |  | ↓ 5 (38%) |  | – / 1 |
| *Well Being (max: 100)* | 8 (4-50) |  | 4 (4-25) |  | ↓ 4 (50%) |  | – / 0 |
| *Stigma (max: 100)* | 0 (0-50) |  | 6 (0-19) |  | – |  | – / 2 |
| *Social support (max: 100)* | 8 (8-33) |  | 17 (0-42) |  | ↑ 9 (100%) |  | – / 1 |
| *Cognition (max: 100)* | 6 (0-19) |  | 19 (0-25) |  | ↑ 13 (200%) |  | – / 3 |
| *Communication (max: 100)* | 17 (8 -25) |  | 25 (8-50) |  | ↑ 8 (50%) |  | – / 3 |
| *Bodily Discomfort (max: 100)* | 17 (8-33) |  | 17 (8-67) |  | 0 (0%) |  | – / 3 |
| MoCA test (max: 30) | 28 (23-29) |  | 28 (22-30) |  | 0 (0%) |  | – |

Minimal Clinically Important Difference (MCID<sup>25</sup>) level for MDS-UPDRS part I is improvement (Imp.) >2.64 points and worsening (Wors.) >2.45 points; for part II is improvement >3.05 points (\*all three patients improved 3.00 points) and worsening >2.51 points; for part III is improvement >3.25 points and worsening >4.63 points; for part IV is improvement >0.9 points and worsening >0.8 points.
\*\*MCID levels for PDQ-39 overall score and subitems are reported only for worsening: PDQ-39 overall score is worsening >1.6 points; for PDQ-39 subitem mobility is worsening > 3.2 points; for PDQ-39 subitem activity of daily living is worsening >4.4 points; for PDQ-39 subitem well-being is worsening >4.2 points; for PDQ-39 subitem stigma is worsening >5.6 points; for PDQ-39 subitem social support is worsening >11.4 points; for PDQ-39 subitem cognition is worsening >1.8; for PDQ-39 subitem communication is worsening >4.2 points; for PDQ-39 subitem bodily discomfort is worsening >2.1 points.
MDS-UPDRS, Movement Disorder Society – Unified Parkinson’s Disease Rating Scale; FoG-Q, Freezing of Gait Questionnaire; MoCA, Montreal Cognitive Assessment; PDQ-39, Parkinson’s Disease Questionnaire - 39 items.

Prior to patient discharge, home monitoring (*Sensing Only*) of beta power and *Events Capture* functions were activated. Patients were instructed to mark specific events *(i*.*e*., *medication intake, bedtime and wake-up time, and, when applicable, troublesome dyskinesia, motor OFF periods and falls)*, for at least three days. The Percept system records a 30-second snapshot of the LFP data immediately following the event. This data is stored on the device and can be reviewed during follow-up visits to identify patterns and correlations between brain activity and patient-reported events.

For the second **ACTIVATION visit** (Table 1, Box 2), patient arrived at the clinic with chronic cDBS active and following overnight withdrawal of antiparkinsonian medication (cDBS-ON/Meds-OFF). During this visit, LFP timelines from the preceding five days were reviewed to assess consistency and variability of the alpha-beta power.

The Timeline function continuously records stimulation parameters, LFP, and events, providing a longitudinal overview of therapy delivery (Figure 2).

**Figure 2.**
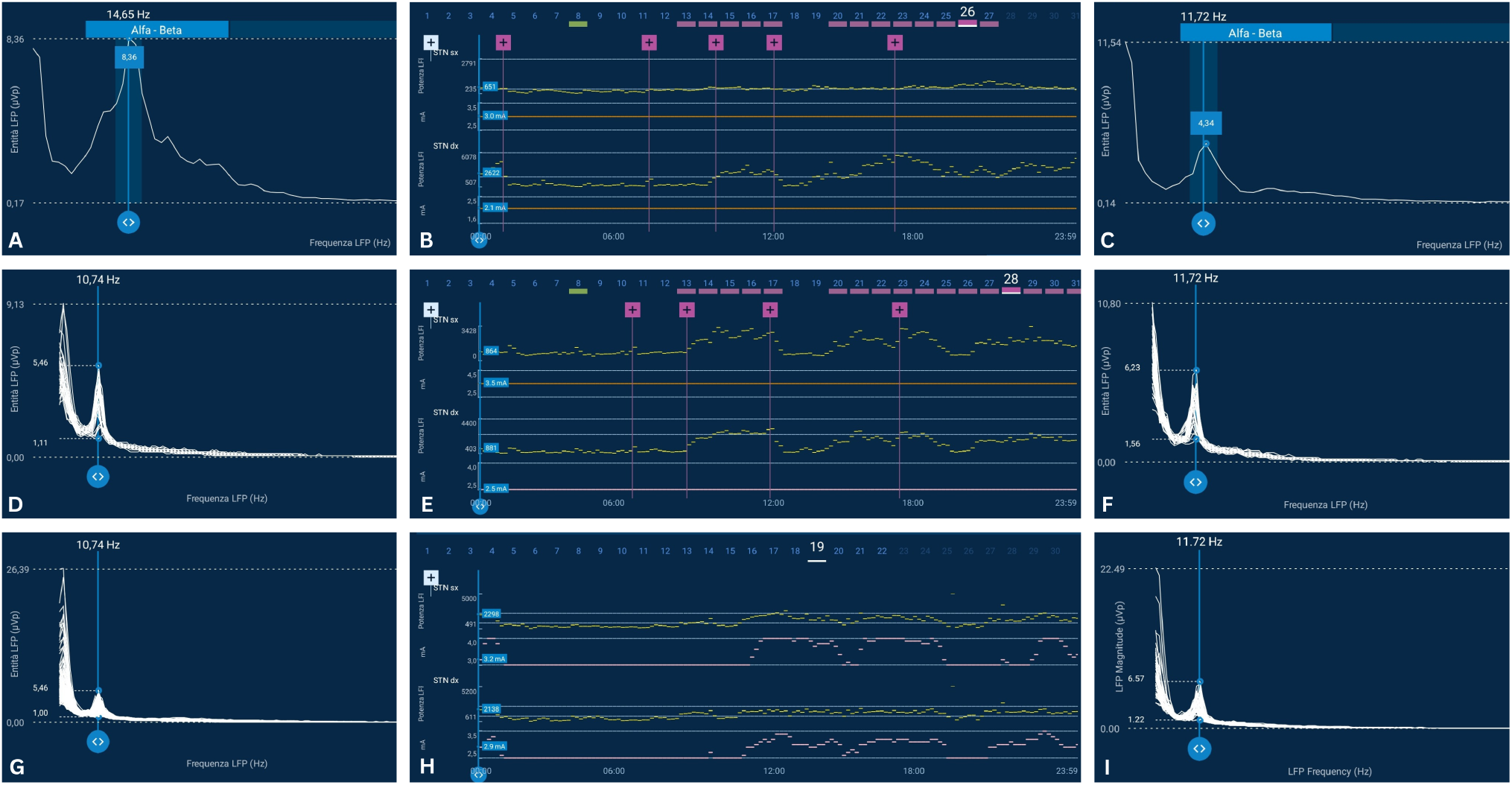
Peak identification with BrainSense Setup, Events LFP Snapshots and corresponding 24-hour modulation of LFP and stimulation. Visualization of the alpha-beta peak of the left STN (14.65 Hz, 8.36 µVp) and right STN (11.72 Hz, 4.34 µVp), identified acutely using *BrainSense Setup* (left STN: A; right STN: C). Its chronic modulation over 24 hours with cDBS (B, left STN upper traces and right STN lower traces) is shown in comparison with the peak (left STN: 10.74 Hz; right STN: 11.72 Hz) identified with *Events LFP Snapshots* (left STN: D and G; right STN: F and I) and its chronic fluctuations over 24 hours during cDBS (E, left STN upper traces and right STN lower traces [for the right STN we activated aDBS, but it resulted in constant stimulation at the lower amplitude limit]) and aDBS (H, left STN upper traces and right STN lower traces). In the central panels (B, E, H), the yellow dashes represent the 10-minute averages of alpha-beta power sampled at 250 Hz within a 5 Hz frequency range centered on the alpha-beta peak. In B and E (left STN), the fixed orange horizontal line indicates cDBS, whereas in panel E (right STN) and H, the pink lines reflect the 10-minute averaged stimulation amplitude delivered over time with aDBS. Notably, in panel E, the left STN was kept on cDBS for a second round of recordings after re-positioning of the alpha-beta patient-specific frequency range due to peak shift. Only the use of the alpha-beta peak defined with *Events LFP Snapshots* enabled the recording of temporal beta fluctuations of the signal in the left STN, which were subsequently used for aDBS programming. For the right STN, it is interesting to note an adequate beta power modulation over 24 hours (the peak selected was at 11.72 Hz and remained unchanged over time), but the thresholds needed to be repositioned: in panel E, the right STN was programmed in aDBS but it resulted in constant stimulated at the lower amplitude limit. Notably, the fluctuation of stimulation amplitude in panel H is present despite beta power modulation remaining between the thresholds (see Discussion).

If confirmed, aDBS using dual thresholds mode was activated as follows:

i. To define LFP thresholds, based on the *Timeline* and *Event LFP Snapshot*, we manually set the lower and upper LFP thresholds at about the 25th and 75th percentiles of beta power recorded during wakeful periods, respectively. We defined these thresholds empirically (visually) as the tablet does not provide a precise calculation of thresholds based on multiple days recordings. Nocturnal beta values –known to decrease during sleep^12,28^– were not considered^5^. Bedtime was identified on patient interviews and the clock of *Timeline* and *Event LFP Snapshot* (Figure 2). Naps during the day were also excluded if referred by the patients. Ramp-up and ramp-down times were kept at the default settings (see above), consistent with those applied in the ADAPT-PD trial^24^. It is worth noting that, in our experience, it is more accurate to identify LFP thresholds based on the chronically recorded data over multiple days rather than the data provided in the clinic over a short period of time by the tablet using the *LFP Thresholds* function. In our experience, with this function, the amplitudes to be used to define LFP thresholds rarely coincide with the clinically selected upper and lower amplitude limits. For the determination of the LFP thresholds, a sufficiently wide gap between amplitudes is required for the system to detect a meaningful difference in beta power at the respective mA levels.
ii. To define stimulation amplitude limits, stimulation was turned off bilaterally, and then (with *BrainSense Streaming*) amplitude was gradually increased in 0.5 mA steps per hemisphere (each hemisphere tested separately, followed by bilateral testing of lower stimulation limit and upper stimulation limit). The lower stimulation limit was defined in the Stim-ON/Meds-OFF condition (overnight pausing of dopaminergic medication), as the amplitude providing >40-50% clinical motor improvement compared to the Stim-OFF/Meds-OFF condition. The upper stimulation limit was defined in the Stim-ON/Meds-ON condition, as the highest effective amplitude prior to the onset of clinical side effects, assessed at least 30 minutes after the intake of the morning dose of antiparkinsonian medications.

A third **FOLLOW-UP visit** for aDBS (Table 1, Box 3) was scheduled about two weeks later to verify patient well-being, effective stimulation modulation, and confirm appropriate placement of thresholds. At the 1-month follow up visit with stable aDBS (unchanged stimulation settings) we repeated the baseline clinical evaluation and questionnaires.

## RESULTS

We evaluated 20 patients (Figure 1): three were implanted with the Medtronic 3389 leads and 17 implanted with the SenSight B33005 directional leads (one with Percept RC). Of these, three patients were excluded due to cardiac artifacts in the LFP signal; two due to activation of the most dorsal contacts in both hemispheres; one due to high stimulation frequency (>185 Hz), which does not allow sensing mode, and one due to psychiatric comorbidities (ideas of reference), which were deemed to compromise the patient’s reliability and compliance with multiple reprogramming sessions.

Of note, the only stimulation-compatible contact pairs with the Percept family devices are the ones that bridge an active contact (see Thenaisie et al.^10^ for details).

LFP artifacts were observed only in patients implanted with Medtronic 3389 electrodes and an IPG placed in the right subclavicular region^29^.

In four patients, the recently introduced *Electrode Identifier* suggested an active contact configuration alternative to that previously clinically defined and used chronically, leading to a decision to reprogram the cDBS settings. As these patients were among the most recently implanted, cDBS optimization was prioritized, and initiation of aDBS was deferred.

As a result, nine patients were deemed eligible for reprogramming in aDBS mode. The study population included three females and six males. The median age was 61 years (range 55-70 years), the median age at symptoms onset was 47 years (range 27-60 years), and the median disease duration was 14 years (range 11-30 years). Additional demographic, clinical and DBS-programming details are reported in Supplementary Materials, Table 1.

Electrodes reconstruction (Supplementary Materials, Figure 1) was obtained with Lead-DBS^30^, confirming accurate targeting of all implanted leads.

At the **SET-UP visit**, five patients showed a bilateral alpha-beta peak during the *BrainSense Surveys* and *BrainSense Setup* and began bilateral recordings. Three patients exhibited an alpha-beta peak in one STN, and recording was activated only for that hemisphere. In two of these patients, one hemisphere could not be assessed because the most dorsal contact was active, making sensing not possible on that hemisphere. One additional patient showed poor (<1.5uVp) alpha-beta peaking in both STNs; nevertheless, we proceeded with bilateral chronic recordings also in this case.

In all patients, the LFP frequency range selected for aDBS programming fell within the alpha-beta band (8-30 Hz).

At the **ACTIVATION visit**, following review of *Timeline* and *Event LFP Snapshots*, the patient-specific alpha-beta frequency range identified during the initial set-up visit was confirmed and maintained in five patients. In two patients, however, a chronic alpha-beta peak was detected in one STN at a frequency different from that observed during the initial BrainSense Signal Test (Figure 2), especially related to “drug intake” events. As a result, sensing-mode recording was repeated for an additional five days, and an extra (fourth) visit was scheduled for these patients. During this extended monitoring period, these patients were programmed with aDBS in one hemisphere and cDBS in the other. At the end of the recording period, sufficient alpha-beta signal was documented, allowing for the initiation of bilateral dual drive aDBS.

At the **FOLLOW-UP visit**, threshold adjustments were required in all patients because the alpha-beta signal either exceeded or fell below the selected thresholds for prolonged periods.

Programming in aDBS mode is still ongoing in three patients and has been deferred to subsequent regular follow-up visits for the following reasons: (i) variability of the alpha-beta peak over time in one patient; and (ii) difficulty in optimizing stimulation parameters with a single drive configuration in two patients. Of note, in single drive configuration, the system allows to adapt stimulation in one hemisphere based on the signal recorded on the other hemisphere.

One patient preferred to return to cDBS. He is a man in his 60’s with a 15-year history of PD, which began with resting tremor in the left upper limb. He has no family history of PD. Approximately six years after diagnosis, he developed motor fluctuations, prolonged OFF states and FOG. Seven years later, he underwent DBS surgery, which resulted in bilateral motor improvement (66% at the MDS-UPDRS part III with best medical treatment before versus after surgery), although FOG persisted. Upon initiation of aDBS, he initially experienced some motor benefit but later developed worsening OFF states. Timeline recordings during follow-up revealed that the alpha-beta signal was frequently below the lower threshold, resulting in under-stimulation. Additional visits were scheduled for this patient at approximately one-week intervals, during which multiple reprogramming strategies were attempted, including adjustments to ramping times (e.g., 10 seconds up, 30 seconds down). It is worth noting, however, that this patient had an atypical cDBS setup, established over time. Specifically, the patient was treated with two distinct stimulation paradigms, which he manually switched between day and night. The two configurations involved different active contacts as well as varying stimulation amplitudes and frequencies (Supplementary Materials, Table 1).

One month after aDBS optimization, the five patients now chronically receiving aDBS underwent the same clinical evaluations performed at the set-up visit (Table 2). All patients showed additional improvement in motor symptoms in the aDBS-ON/Meds-OFF condition compared to cDBS-ON/Meds-OFF, with a 35% improvement in MDS-UPDRS III scores and a 40% improvement in FOG-Q scores. The improvement in MDS-UPDRS III scores reached the MCID^25^ level in four of the five patients (Table 2). Of note, we cannot exclude that the additional benefit reflected in the MDS-UPDRS III scores may partly relate to the patient’s medication state (i.e., overnight Meds-OFF) at the time of the visit, which can be associated with greater beta power and, consequently, higher stimulation amplitudes in aDBS compared with cDBS.

Regarding non-motor symptoms, MDS-UPDRS parts I and II improved by 18% and 11%, respectively, at the MCID level in three patients (Table 2). However, these improvements were not reflected in the PDQ-39 total score, which on average worsened by 28% under aDBS (with three patients reaching the MCID level)^31^. Nonetheless, analysis of PDQ-39 sub-scores showed improvements in both Activities of Daily Living and Well-Being (Table 2).

All patients who were able to use aDBS chronically, and their caregivers, expressed a preference for aDBS over conventional cDBS and continue to receive aDBS therapy.

The average total electrical energy delivered (TEED) in cDBS was 46.8µW in the left STN and 39.3µW in the right STN, whereas in aDBS, it was 53.0µW and 48.5µW, respectively. On average, we delivered 12% more TEED in the left STN and 19% more in the left STN with aDBS compared to cDBS. TEED was calculated for aDBS over a median period of 35 days (range: 12–43 days) (Supplementary Materials, Table 2).

The amplitude reported for both aDBS and cDBS, and used to calculate TEED, corresponds to the “display amplitude” – a single value, approximately three times the stimulation amplitude from the strongest segment, and not the “delivered amplitude”, which represents the sum of stimulation amplitudes delivered by each active electrode. The aDBS summary uses the “display amplitude”^32^.

## DISCUSSION

Our initial experience in a limited, non-blinded sample with relatively short follow-up suggests that aDBS in dual threshold mode may further enhance the benefit achieved with chronically optimized cDBS in selected patients. This observation is particularly noteworthy given that cDBS had already provided substantial clinical benefit in our cohort (Table 2).

The additional improvement achieved with the aDBS mode was documented through clinical neurological assessment. However, this approach may be suboptimal, as it might not fully capture the patient’s experience over time in their daily environment, where aDBS is hypothesized to be more effective due to its capacity to dynamically adjust stimulation amplitude as needed.

To better assess the real-world impact of aDBS, patients completed the FOG-Q and PDQ-39 referencing the previous weeks. Surprisingly, PDQ-39 total scores worsened under aDBS compared with cDBS, despite improvements in FOG-Q and MDS-UPDRS scores and a clear preference for aDBS reported by patients and caregivers. To account for this discrepancy, we hypothesize that patients’ increased engagement in daily activities –possibly due to improved gait and motor function with aDBS (as indicated by FOG-Q and MDS-UPDRS scores)– may have introduced new challenges during the limited assessment period. A reduction in caregiver support, resulting from improvement in parkinsonian symptoms, could also have contributed to the worsening of the PDQ-39 Social Support subscore. Furthermore, the reported worsening of PDQ-39 Cognition and Communication subscores was not confirmed by MoCA testing, which is considered more objective than patient self-reports and is consistent with preliminary evidence supporting the cognitive safety of aDBS^33^.

In any case, patient preference –particularly when aligned with caregiver input and clinical judgment– should, in our view, be considered the most relevant outcome. These findings suggest caution in relying solely on retrospective self-report questionnaires when evaluating the long-term effects of different DBS modes. Eventually, patients should also be instructed and trained to complete these questionnaires by specifically recalling and comparing different clinical conditions. The integration of digital diaries for real-time data collection, combined with wearable sensors monitoring (currently under analysis), could provide more accurate and reliable assessments of patient outcomes. This aspect is particularly important given the strong placebo effect in patients with PD, especially in longitudinal open-label studies of new therapeutic strategies that raise high expectations^20,22,34^.

It is important to note that the decision not to adjust the therapeutic regimen –specifically, the number and dosage of daily levodopa intake– may have disadvantaged the aDBS condition, as the current algorithms are designed to modulate stimulation based on levodopa-induced fluctuations in the beta power biomarker and, therefore, may better allow optimization of medication.

In our study, the average TEED was higher in aDBS compared to cDBS. However, the delivered current was not continuously and consistently higher. This is a key advantage of aDBS, as it automatically delivers higher current only when necessary. A direct comparison of the TEED between cDBS and aDBS was feasible only as averages, which are automatically calculated by the system and reported in the *Summary* of the stimulation period (Supplementary Materials, Table 2).

It is worth noting that, for aDBS programming, we intended to maximize the gap between the lower and the upper stimulation limits to fully exploit the advantages of current modulation, aiming for an empirical minimum difference of 0.7 mA between the two amplitude limits. However, this target could not be achieved in one patient (Patient 5; Supplementary Materials, Table 1), for whom a slightly smaller gap (0.6 mA bilaterally) was used instead in a single drive configuration.

In two patients (Patients 1 and 3; Supplementary Materials, Table 1), the lower stimulation limit corresponded to –or even exceeded– the amplitude used during cDBS. This configuration was necessary to maintain consistent tremor suppression, as tremor tended to re-emerge at lower stimulation levels than those applied in cDBS. We hypothesize that this may reflect a habituation effect, possibly requiring a longer period to reverse; however, further studies are needed to elucidate this aspect^35^.

Despite the promising clinical performance of aDBS, several technical limitations were encountered with the dual threshold mode, restricting its applicability to approximately half of the patients assessed. Below, we outline the specific challenges faced during each phase of the programming process.

As for the ADAPT-PD study, patients were asked to attend the **SET-UP visit** after an overnight withdrawal of all dopaminergic medications. In our case, in addition to providing a baseline clinical state for comparison between cDBS and aDBS, medication withdrawal was necessary to optimize alpha-beta peak detection, as beta power is known to be suppressed by levodopa^7^. Medication withdrawal can be challenging for patients, even with active stimulation. In the future, end-of-dose assessments may offer a more feasible alternative for evaluating motor symptoms and local field potentials, and determining the lower stimulation limit, although this approach would require careful scheduling of outpatient visits.

In some patients, despite being in the Stim-OFF/Meds-OFF condition, manual identification of the alpha-beta peak during the *BrainSense Setup* was required due to the system inability to detect it automatically. Poor alpha-beta peak identification was also observed in patients implanted with SenSight leads. In such cases, real-time LFP streaming was essential to confirm beta signal modulation in response to stimulation. Artifact contamination of the LFP remains a significant limitation of Percept devices, leading to the exclusion of three patients implanted with Medtronic 3389 leads from aDBS activation.

In our study, we excluded patients with LFPs contaminated by cardiac or gait-related artifacts. At present, we recommend this approach, as it remains unclear which parameters (e.g., onset time, onset duration) can be adjusted to mitigate their impact on aDBS programming^29,36^. In our view, a visual assessment of artifact presence, together with the information provided by the tablet, may be sufficient to exclude contacts with contaminated recordings. More detailed evaluations, whether clinical or instrumental (e.g., ECG recordings or gait analysis), could increase accuracy but substantially prolong the duration of the visit.

Activation of the sensing mode requires the selection of sensing contacts that bridge the stimulation contacts, and leads of the directional level cannot be used simultaneously for both sensing and stimulation. This technical constraint limits contact configuration and, in our cohort, excluded two patients from aDBS activation. This remains one of the principal limitations of the Percept device, as we currently advise against selecting stimulating contacts based on the feasibility of applying aDBS paradigms rather than on clinical efficacy. This consideration should be integrated into surgical planning to ensure accurate placement of the two central (segmented) contacts at the intended target. Additionally, sensing can be enabled only when stimulation frequency is in the range 55-180Hz (SenSight leads) or 50-185Hz (8839 leads), a limitation that prevented aDBS activation in one patient.

Lastly, the automated threshold suggestion does not consider the broader day-to-day variability in beta activity seen in chronic recordings, leading to suboptimal settings.

The primary aim of the **ACTIVATION visit** was to review five days of *BrainSense Timeline* and *Event LFP Snapshots* recordings to confirm the reliability of the LFP signal observed during the set-up visit. Initial aDBS programming could be attempted during this visit; however, follow-up assessments were necessary for fine-tuning.

We found that home-based LFP monitoring is critical for optimal aDBS programming. In two patients, the frequency range required repositioning based on *Event LFP Snapshots*, necessitating an additional five days of sensing-mode recording. Notably, home monitoring at this stage was performed with active cDBS, and follow-up after aDBS activation remained essential for further reprogramming. This iterative process proved particularly relevant given the narrow 5Hz frequency window allowed by the Percept for biomarker detection. Even minor shifts in the selected peak frequency could significantly impair aDBS performance (Figure 2). The cause of discrepancies between acutely and chronically recorded alpha-beta peak frequencies –despite being only five days apart within the same patient– remains unclear and warrants further investigation.

We chose a five-day recording period; however, based on our experience, a three-day period may still be sufficient to reliably assess circadian beta variability, its association with akinetic-rigid symptoms, and its responsiveness to dopaminergic therapy –particularly levodopa. Nevertheless, because large LFP outliers may complicate *BrainSense Timeline* interpretation, we generally recommend extended home recordings, which allow for the exclusion of days with artifacted or non-representative LFP data. In contrast, a single in-clinic recording session, as proposed in the ADAPT-PD study^24^, appears insufficient for the dual threshold aDBS mode.

When the signal was available from only one hemisphere, aDBS was initiated in single drive mode, with stimulation adjustments applied bilaterally based on unilateral recordings. This approach was based on previous studies indicating that beta peak frequencies and their fluctuations are generally similar between left and right STNs in most PD patients^37^. Future research should further evaluate these findings and systematically compare single drive versus dual drive aDBS.

Contrary to previous reports^20^, we set the lower and upper stimulation limits using a conservative approach, ensuring that patients never fell below 40% of therapeutic efficacy (lower stimulation limit/Meds-OFF) and experienced no side effects at the upper stimulation limit/Meds-ON (peak levodopa effect) (see Methods – Activation Visit). With this approach, in half of the STNs the gap between limits exceeded 1 mA, remaining free of adverse effects during follow-up. No patients reported troublesome dyskinesias, OFF states, or falls, and although permitted, none required a switch from aDBS back to cDBS during follow-up. This programming strategy was informed by the counterintuitive observation that, in some patients, elevated beta power sometimes persisted despite clear clinical benefit during peak-dose levodopa, whereas in others, beta power remained low even in a clinically defined end-of-dose state. These findings underscore the need for further studies investigating beta responsiveness to levodopa, particularly under chronic aDBS.

It should also be noted that the dual threshold algorithm maintains a constant stimulation amplitude equal to the last value reached when the LFP signal remains between the two thresholds, which may result in prolonged periods at either the upper or lower stimulation limit before any adjustment occurs. For example (see Figure 3 of the SSED^38^), if a patient is at the upper stimulation limit due to elevated LFP activity (above the upper threshold) and the signal subsequently decreases into the intermediate range, approaching but not crossing the lower threshold, the stimulation amplitude remains fixed at the upper limit despite the patient’s potential clinical need for reduced stimulation.

A technical limitation also emerged during the definition of LFP thresholds. The *BrainSense Timeline* displays beta power as 10-minute averages, thereby masking real fluctuations. This poor temporal resolution may compromise optimal threshold selection, as the dynamic properties of the beta signal are not displayed.

During **FOLLOW-UP visits** under chronic aDBS, threshold adjustments were necessary due to variability in beta-band signal power. In some cases, analysis of the *BrainSense Timeline* showed that stimulation alternated between lower and upper stimulation limits only a few times throughout the day, resulting in a two-level amplitude cDBS pattern rather than true aDBS modulation.

Establishing thresholds based on 10-minute averaged beta values proved unreliable, while changes in stimulation amplitude over time may help guide threshold adjustments. This is illustrated in Figure 2 (Box F), where beta power fluctuations between thresholds would be expected to result in no change in stimulation amplitude; instead, a marked adaptation of the delivered stimulation within the defined limits is observed. This approach remains suboptimal, as amplitude is a programmable output rather than a direct reflection of the underlying biomarker (i.e., the brain signal). The low time resolution (10-minute averages) should be improved to provide a clearer visualization of the actual fluctuations in LFP and stimulation amplitude.

The need for repeated threshold optimization in some patients may increase the number of unscheduled clinical visits required to achieve effective aDBS performance. Postponing the initiation of aDBS to subsequent outpatient visits, as in our case, is feasible for the SET-UP visit; however, the ACTIVATION and FOLLOW-UP visits should remain relatively close in time to allow for timely adjustment of stimulation parameters. These challenges underscore the importance of implementing remote monitoring and programming capabilities to enable more efficient and individualized patient management.

In conclusion, aDBS appears safe, well tolerated, and preferred by patients over cDBS. It shows promise for symptoms like gait impairment that are suboptimally controlled with cDBS. However, optimizing dual threshold parameters remains challenging and requires validation in larger cohorts. Identifying reliable biomarkers, improving automated programming^39^, and refining algorithms are essential to advancing chronic aDBS in clinical practice. Nevertheless, contrary to previous aDBS programming strategies using the dual threshold algorithm with the Percept device^20,38^, the proposed protocol allowed us to meet our main objectives, which, in addition to providing greater clinical benefit,remained avoiding troublesome adverse effects (none reported) and maintaining a low number of additional visits (limited to 3-4 appointments) for aDBS initiation.

## Funding

The study was funded by the Fondazione Pezzoli per la Malattia di Parkinson and the European Union–Next-Generation EU - NRRP M6C2 - Investment 2.1 Enhancement and strengthening of biomedical research in the NHS (Project-ID PNRR-MAD-2022-12376927). FL, CP and IUI were supported by the Deutsche Forschungsgemeinschaft (DFG, German Research Foundation) Project-ID 424778381 - TRR 295. FL and LC were supported by a grant of the German Excellence Initiative to the Graduate School of Life Sciences, University of Würzburg. IUI was supported by a grant from New York University School of Medicine and The Marlene and Paolo Fresco Institute for Parkinson’s and Movement Disorders, which was made possible with support from Marlene and Paolo Fresco.

The funders played no role in study design, data collection, analysis and interpretation of data, or the writing of this manuscript.

## Institutional Review Board Statement

This study was conducted in accordance with the Declaration of Helsinki and approved by the institutional review board of Comitato Etico Milano Area 2 and Comitato Etico Territorial Lombardia 3.

## Informed Consent Statement

All patients provided written informed consent.

## Acknowledgements

We are grateful to Philipp Capetian, Nicoló G. Pozzi, Ibrahem Hanafi and Aya Al Habbal of the University Hospital of Würzburg for critically reading the manuscript and to Rebecca Barbiani and Ilaria Riela of the Fondazione Pezzoli for Parkinson’s Disease and the ASST G.Pini-CTO for administrative support. We are also thankful to Gianluca Cassano, Federico Cavallini, Gaetano Leogrande, Lucia Limido, Valentina Di Russo and Scott Stanslaski of Medtronic Inc. for their valuable comments on the correctness and the clarity of the device-related technical details of the manuscript. Medtronic Inc. played no role in study design, data collection, analysis and interpretation of data, or the writing of this manuscript.

## CRediT – Contributor Roles Taxonomy

Conceptualization: SC, FL, EC, IUI, SB

Methodology: SC, FL, EC, LC, CP, IUI, SB

Formal Analysis: SC, FL, LC, CP, SB

Investigation: SC, FL, EC, LC, CP, IUI, SB

Data Curation: SC, FL, LC, CP

Validation: EC, CP, IUI, SB

Resources: GP, IUI

Writing: SC, FL, SB

Writing – Review & Editing: EC, LC, CP, GP, IUI

Visualization: SC, FL, EC, SB

Supervision: IUI, SB

Project Administration: IUI

Funding Acquisition: CP, GP, IUI

## Competing interests

Author IUI received lecture honoraria and research fundings from Medtronic Inc. and Boston Scientific, but declares no financial or non-financial competing interests. Author IUI is consultant, holds share [Newronika S.p.A.] and received funding for research activities from Newronika S.p.A., but declares no financial or non-financial competing interests. Author IUI serves as Adjunct Professor at the NYU Grossman School of Medicine and the University of Milan, but declares no financial or non-financial competing interests.

Author CP received research funding from Medtronic Inc., but declares no financial or non-financial competing interests. All other authors declare no financial or non-financial competing interests.

## Data availability

The datasets generated and/or analysed during the current study are not publicly available due to privacy law but are available from the corresponding author on reasonable request.

## Notes

### Competing Interest Statement

The authors have declared no competing interest.

## REFERENCES

1. Aum DJ, Tierney TS. Deep brain stimulation foundations and future trends. Front Biosci 2018;23(1):162–182.

2. St. George RJ, Nutt JG, Burchiel KJ, Horak FB. A meta-regression of the long-term effects of deep brain stimulation on balance and gait in PD. Neurology 2010;75(14):1292–1299.

3. Oehrn CR, Cernera S, Hammer LH, et al. Chronic adaptive deep brain stimulation versus conventional stimulation in Parkinson’s disease: a blinded randomized feasibility trial. Nat Med 2024;30(11):3345–3356.

4. Little S, Pogosyan A, Neal S, et al. Adaptive deep brain stimulation in advanced Parkinson disease. Annals of Neurology 2013;74(3):449–457.

5. Gobeil M, Guillemette A, Silhadi M, et al. Local Field Potential Biomarkers of Non‐Motor Symptoms in Parkinson’s Disease: Insights From the Subthalamic Nucleus in Deep Brain Stimulation. Eur J of Neuroscience 2025;61(5):e70046.

6. Neumann W, Degen K, Schneider G, et al. Subthalamic synchronized oscillatory activity correlates with motor impairment in patients with Parkinson’s disease. Movement Disorders 2016;31(11):1748–1751.

7. Kühn AA, Kupsch A, Schneider G, Brown P. Reduction in subthalamic 8–35 Hz oscillatory activity correlates with clinical improvement in Parkinson’s disease. Eur J of Neuroscience 2006;23(7):1956–1960.

8. Priori A, Foffani G, Rossi L, Marceglia S. Adaptive deep brain stimulation (aDBS) controlled by local field potential oscillations. Experimental Neurology 2013;245:77–86.

9. Kuhn AA, Kempf F, Brucke C, et al. High-Frequency Stimulation of the Subthalamic Nucleus Suppresses Oscillatory Activity in Patients with Parkinson’s Disease in Parallel with Improvement in Motor Performance. Journal of Neuroscience 2008;28(24):6165–6173.

10. Thenaisie Y, Palmisano C, Canessa A, et al. Towards adaptive deep brain stimulation: clinical and technical notes on a novel commercial device for chronic brain sensing. J. Neural Eng. 2021;18(4):042002.

11. Muller M, Scafa S, Hanafi I, et al. Online prediction of optimal deep brain stimulation contacts from local field potentials in Parkinson’s disease. npj Parkinsons Dis. 2025;11(1):234.

12. Caffi L, Romito LM, Palmisano C, et al. Adaptive vs. Conventional Deep Brain Stimulation: One-Year Subthalamic Recordings and Clinical Monitoring in a Patient with Parkinson’s Disease. Bioengineering 2024;11(10):990.

13. Avantaggiato F, Farokhniaee A, Bandini A, et al. Intelligibility of speech in Parkinson’s disease relies on anatomically segregated subthalamic beta oscillations. Neurobiology of Disease 2023;185:106239.

14. Vissani M, Palmisano C, Volkmann J, et al. Impaired reach-to-grasp kinematics in parkinsonian patients relates to dopamine-dependent, subthalamic beta bursts. npj Parkinsons Dis. 2021;7(1):53.

15. Canessa A, Palmisano C, Isaias IU, Mazzoni A. Gait-related frequency modulation of beta oscillatory activity in the subthalamic nucleus of parkinsonian patients. Brain Stimulation 2020;13(6):1743–1752.

16. Arnulfo G, Pozzi NG, Palmisano C, et al. Phase matters: A role for the subthalamic network during gait. PLoS ONE 2018;13(6):e0198691.

17. Neumann W, Gilron R, Little S, Tinkhauser G. Adaptive Deep Brain Stimulation: From Experimental Evidence Toward Practical Implementation. Movement Disorders 2023;38(6):937–948.

18. Bocci T, Prenassi M, Arlotti M, et al. Author Correction: Eight-hours conventional versus adaptive deep brain stimulation of the subthalamic nucleus in Parkinson’s disease. npj Parkinsons Dis. 2022;8(1):18.

19. Isaias IU, Caffi L, Borellini L, et al. Case report: Improvement of gait with adaptive deep brain stimulation in a patient with Parkinson’s disease. Front. Bioeng. Biotechnol. 2024;12:1428189.

20. Busch JL, Kaplan J, Behnke JK, et al. Chronic adaptive deep brain stimulation for Parkinson’s disease: clinical outcomes and programming strategies. npj Parkinsons Dis. 2025;11(1):264.

21. Isaias IU, Marceglia S, Borellini L, et al. Chronic adaptive versus conventional deep brain stimulation in Parkinson’s disease: a blinded randomized pilot trial. medRxiv 2025;2025.02.20.25322374.

22. Bronte-Stewart HM, Beudel M, Ostrem JL, et al. Long-Term Personalized Adaptive Deep Brain Stimulation in Parkinson Disease: A Nonrandomized Clinical Trial. JAMA Neurol 2025;e252781.

23. Montgomery EB. Introduction [Internet]. Oxford University Press; 2016.[cited 2025 Aug 6] Available from: https://academic.oup.com/book/24740/chapter/188236572

24. Stanslaski S, Summers RLS, Tonder L, et al. Sensing data and methodology from the Adaptive DBS Algorithm for Personalized Therapy in Parkinson’s Disease (ADAPT-PD) clinical trial. npj Parkinsons Dis. 2024;10(1):174.

25. Mishra B, Sudheer P, Rajan R, et al. Bridging the gap between statistical significance and clinical relevance: A systematic review of minimum clinically important difference (MCID) thresholds of scales reported in movement disorders research. Heliyon 2024;10(5):e26479.

26. Klocke P, Loeffler MA, Lewis SJG, et al. Could adaptive deep brain stimulation treat freezing of gait in Parkinson’s disease? J Neurol 2025;272(4):267.

27. Galeoto G, Colalelli F, Massai P, et al. Quality of life in Parkinson’s disease: Italian validation of the Parkinson’s Disease Questionnaire (PDQ-39-IT). Neurol Sci 2018;39(11):1903–1909.

28. Van Rheede JJ, Feldmann LK, Busch JL, et al. Diurnal modulation of subthalamic beta oscillatory power in Parkinson’s disease patients during deep brain stimulation. npj Parkinsons Dis. 2022;8(1):88.

29. Neumann W-J, Memarian Sorkhabi M, Benjaber M, et al. The sensitivity of ECG contamination to surgical implantation site in brain computer interfaces. Brain Stimulation 2021;14(5):1301–1306.

30. Neudorfer C, Butenko K, Oxenford S, et al. Lead-DBS v3.0: Mapping deep brain stimulation effects to local anatomy and global networks. NeuroImage 2023;268:119862.

31. Peto V. Determining minimally important differences for the PDQ-39 Parkinson’s disease questionnaire. Age and Ageing 2001;30(4):299–302.

32. Model B35200 and Model B35300 Percept− neurostimulators with BrainSense− technology and Adaptive Therapy [Internet]. [date unknown];[cited 2025 Sept 24] Available from: https://www.accessdata.fda.gov/cdrh_docs/pdf/P960009S478C.pdf

33. Ferrucci R, Ruggiero F, Aiello EN, et al. Cognitive effects of adaptive deep brain stimulation in Parkinson’s disease: stability without risk. Eur J Med Res 2025;30(1):820.

34. Lidstone SC, Schulzer M, Dinelle K, et al. Effects of Expectation on Placebo-Induced Dopamine Release in Parkinson Disease. Arch Gen Psychiatry 2010;67(8):857.

35. Fasano A, Helmich RC. Tremor habituation to deep brain stimulation: Underlying mechanisms and solutions. Movement Disorders 2019;34(12):1761–1773.

36. Sousa M, Tinkhauser G. Being Guided by Your Brain or by Your Heart? Challenges in Adaptive Deep Brain Stimulation. Movement Disord Clin Pract 2025;mdc3.70337.

37. De Solages C, Hill BC, Koop MM, et al. Bilateral symmetry and coherence of subthalamic nuclei beta band activity in Parkinson’s disease. Experimental Neurology 2010;221(1):260–266.

38. https://www.accessdata.fda.gov/cdrh_docs/pdf/P960009S478B.pdf. [date unknown];

39. Falciglia S, Caffi L, Baiata C, et al. Transformer-based long-term predictor of subthalamic beta activity in Parkinson’s disease. npj Parkinsons Dis. 2025;11(1):210.

