## Supplementary Materials for "Chronic Adaptive Deep Brain Stimulation in Parkinson’s Disease: ADAPT-START Findings and Programming Principles"

**Supplementary Figure 1 – Localization, reconstruction, and visualization of the implanted electrodes in patients who successfully underwent aDBS follow-up.**

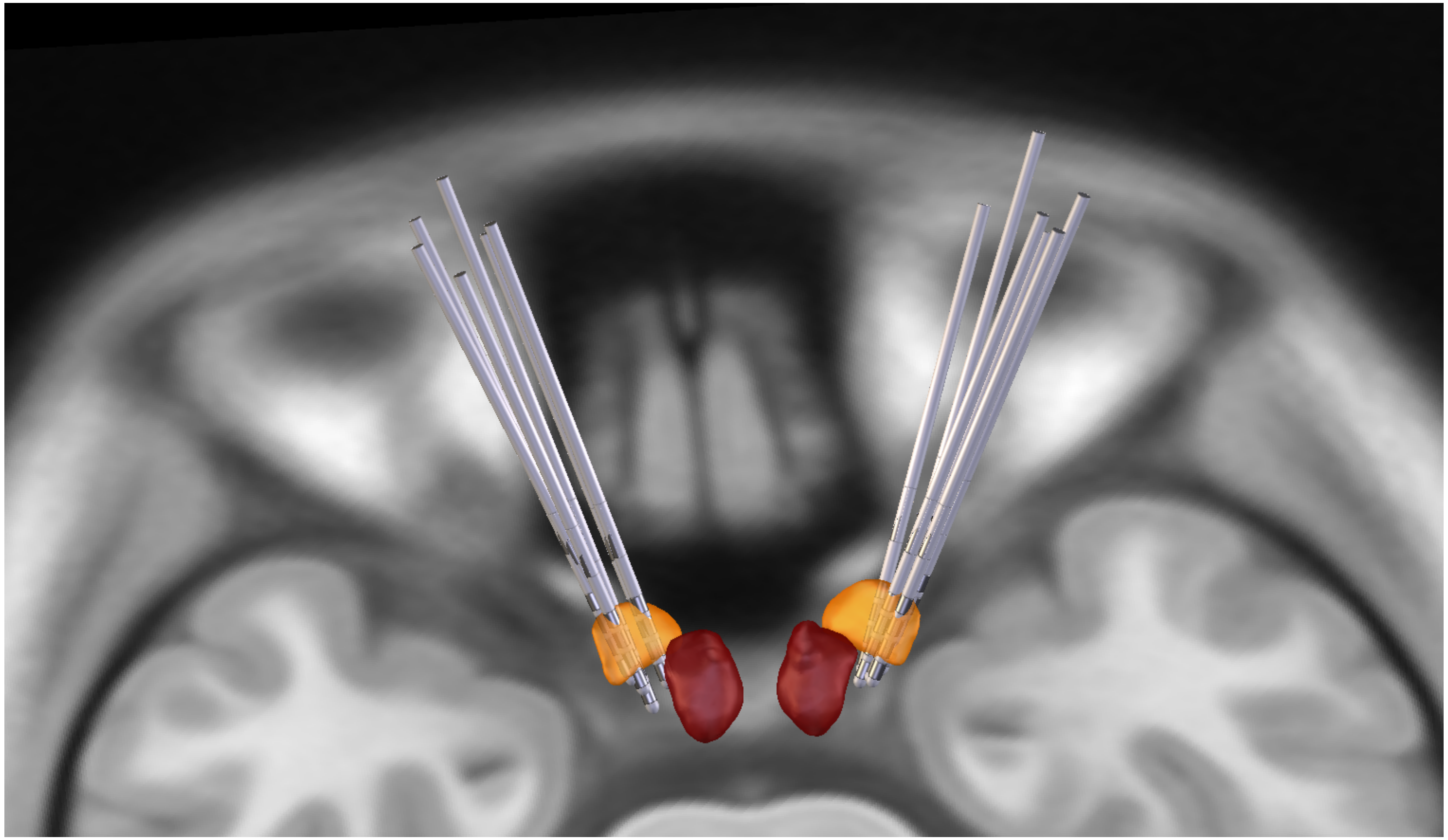

We employed Lead Group Analysis (Lead-DBS v3.2) and the DISTAL Minimal subcortical atlas (Ewert et al., 2017[30]) for electrode localization, reconstruction, and visualization. The subthalamic nucleus (STN) is highlighted in orange, while the red nucleus is shown in red (https://www.lead-dbs.org/). This figure confirms the precise targeting of all implanted leads.

### Supplementary Table 1 – Demographic and clinical features

| Patient | Gender (M/F) | Age (range) | Age at onset (range) | Disease duration (year) | Duration of cDBS therapy (month) | Medications | LEDD (mg) | MDS-UPDRS III cDBS-OFF / Meds-OFF (score) | cDBS parameters[#] | | LFP peak (Hz) | | aDBS amplitude parameters (mA) | | | | | |
|---|---|---|---|---|---|---|---|---|---|---|---|---|---|---|---|---|---|---|
| | | | | | | | | | Left STN | Right STN | Left STN | Right STN | Left STN | | | Right STN | | |
| | | | | | | | | | | | | | Stimulation amplitude limits | | | Stimulation amplitude limits | | |
| | | | | | | | | | | | | | Lower | Pause | Upper | Lower | Pause | Upper |
| 1 | M | 66-70 | 51-55 | 11 | 12 | Levodopa/Carbidopa 100/25 mg:<br>1/2 cp three times/day | 150 | 33 | C+ 2-<br>3 mA<br>60 µs<br>125 Hz | C+ 10b-10c-<br>2.1 mA<br>60 µs<br>125 Hz | 10.74 | 11.72 | 3 | 3.5 | 4 | 2.5 | 3 | 3.5 |
| 2 | F | 66-70 | 51-55 | 11 | 11 | Levodopa/Benserazide 200/50 mg:<br>1/2 cp three times/day | 300 | 58 | C+ 2-<br>2.6 mA<br>60 µs<br>125 Hz | C+ 10a-10b-<br>2.4 mA<br>50 µs<br>125 Hz | 12.70 | 12.70 | 2.3 | 2.7 | 3.5 | 2.2 | 2.5 | 3.2 |
| 3 | M | 61-65 | 36-40 | 24 | 9 | Levodopa/Carbidopa 100/25 mg:<br>1 cp four times/day<br>Pramipexole 0,7 mg:<br>1 cp three times/day | 610 | 44 | C+ 2a-2b-<br>2.6 mA<br>60 µs<br>125 Hz | C+ 9-<br>2.4 mA<br>60 µs<br>125 Hz | 22.46 | 22.46 | 2.7 | 2.9 | 3.4 | 2.7 | 2.9 | 3.5 |
| 4 | F | 56-60 | 26-30 | 30 | 13 | Levodopa/Benserazide 200/50 mg:<br>1/4 cp three times/day;<br>Ropinirole 8 mg RP:<br>1 cp/day | 310 | 33 | C+ 2a-2b-<br>2.5 mA<br>60 µs<br>125 Hz | C+ 9a-9b-<br>3 mA<br>60 µs<br>125 Hz | 17.58 | 21.48 | 2.2 | 2.5 | 3 | 2.5 | 3 | 3.5 |
| 5 | F | 56-60 | 41-45 | 17 | 9 | Levodopa/Benserazide 100/25 mg:<br>1+1/2 cp once/day,<br>1 cp three times/day<br>Ropinirole 4 mg RP:<br>1 cp/day | 530 | 37 | C+ 2-<br>2.3 mA<br>40 µs<br>85 Hz | C+ 10-<br>1.8 mA<br>60 µs<br>85 Hz | 23.44 | // | * 2.1 | 2.3 | 2.7 | 1.6 | 1.8 | 2.1 |
| 6 | M | 51-55 | 46-50 | 8 | 10 | // | // | 34 | C+ 2a- 2c-<br>2.9 mA<br>60 µs<br>180 Hz | 10+11-<br>1.8 mA<br>60 µs<br>180 Hz | // | | Still optimizing aDBS | | | // | | |
| 7 | M | 66-70 | 56-60 | 9 | 6 | Levodopa/Carbidopa 100/25 mg:<br>1+1/2 cp twice/day,<br>1 cp three times/day<br>Rotigotine patch 8 mg/day | 840 | 48 | C+ 1a-2a-<br>3.5 mA<br>60 µs<br>125 Hz | C+ 10c-<br>3.6 mA<br>60 µs<br>125 Hz | // | | Still optimizing aDBS | | | // | | |
| 8 | M | 66-70 | 56-60 | 14 | 7 | Melevodopa/Carbidopa 25mg+100mg:<br>1 cp five times/day<br>Pramipexole RP 1.05 mg:<br>1 cp/day | 605 | 61 | C+ 2b-2c-<br>4.5 mA<br>50 µs<br>180 Hz | C+ 10-<br>2.3 mA<br>60 µs<br>180 Hz | // | | Still optimizing aDBS | | | // | | |

* STN single drive aDBS

\# The amplitude reported for both aDBS and cDBS corresponds to the "display amplitude" – a single value, approximately three times the stimulation amplitude from the strongest segment – and not the "delivered amplitude", which represents the sum of stimulation amplitudes delivered by each active electrode. The aDBS summary uses the "display amplitude".

### Supplementary Table 2 – Total Electrical Energy Delivered in cDBS and aDBS

| Patient | TEED cDBS (μW) | | TEED aDBS (μW) | | aDBS parameters | | | | | | | | | | | | | |
|---|---|---|---|---|---|---|---|---|---|---|---|---|---|---|---|---|---|---|
| | | | | | Left STN | | | | | | | Right STN | | | | | | |
| | Left STN | Right STN | Left STN | Right STN | Average amplitude (mA) | Upper LFP Threshold (a.u.) | Lower LFP Threshold (a.u.) | Above Threshold (%) | Between Threshold (%) | Below Threshold (%) | Days in aDBS for TEED calculation* | Average amplitude (mA) | Upper LFP Threshold (a.u.) | Lower LFP Threshold (a.u.) | Above Threshold (%) | Between Threshold (%) | Below Threshold (%) | Days in aDBS for TEED calculation a |
| 1 | 67.5 | 33.1 | 86.2 | 56.7 | 3.39 | 2500 | 1200 | 5 | 54 | 40 | 42 | 2.75 | 2600 | 1300 | 1 | 67 | 32 | 42 |
| 2 | 50.7 | 36.0 | 58.4 | 42.0 | 2.79 | 3600 | 2070 | 4 | 84 | 13 | 43 | 2.59 | 2700 | 1200 | 4 | 69 | 27 | 43 |
| 3 | 50.7 | 43.2 | 66.2 | 62.6 | 2.97 | 450 | 250 | 23 | 50 | 25 | 35 | 2.89 | 620 | 350 | 11 | 45 | 42 | 35 |
| 4 | 46.9 | 67.5 | 36.0 | 65.2 | 2.19 | 1150 | 750 | 3 | 46 | 51 | 43 | 2.95 | 150 | 100 | 19 | 57 | 23 | 43 |
| 5 | 18.0 | 16.5 | 18.0 | 16.0 | 2.30 | 240 | 150 | 14 | 54 | 32 | 31 | 1.77 | // | // | 14 | 54 | 32 | 31 |

* These are the days used to calculate the Total Electrical Energy Delivered (TEED) in optimized aDBS mode, i.e., without any programming adjustments, and include the follow-up month. The TEED for both cDBS and aDBS was calculated using the "display amplitude"[32]. The aDBS summary uses the "display amplitude".
